# Biallelic *PARP1* Mutations Associated with Childhood-Onset Neurodegeneration

**DOI:** 10.1101/2023.06.09.23291078

**Authors:** Richard Hailstone, Reza Maroofian, Lisa Woodbine, Elena Korneeva, Jan Brazina, Alfons Macaya, Mariasavina Severino, Hoda Tomoum, Henry Houlden, Keith W Caldecott

## Abstract

PARP1 is the primary human sensor protein for DNA single-strand breaks, reduced repair of which results in neurodevelopmental and/or progressive neurodegenerative disease typified by cerebellar ataxia, and oculomotor apraxia. Here, we report the first such disease associated with hereditary mutations in *PARP1*. The affected individual possesses biallelic mutations in the second DNA strand-break sensing zinc finger of PARP1, resulting in a predicted truncated protein of 127 amino acids that is comprised of just the first DNA strand-break sensing zinc finger. Levels of oxidative DNA damage-induced PARP activity are greatly reduced (∼80%) in primary fibroblasts derived from the patient, when compared to cells derived from the parents, and levels of endogenous S-Phase PARP activity are reduced by >50%. Critically, whereas DNA double-strand break repair and cellular sensitivity to ionising radiation are largely normal in the patient-derived cells, the rate of repair of DNA single-strand breaks induced by either oxidative stress, during DNA base excision repair, or as a result of cytotoxic topoisomerase I activity is reduced. These data implicate hereditary mutations PARP1 in human hereditary neurodegenerative disease, and increase to five the number of DNA single-strand break repair genes associated with progressive cerebellar ataxia and oculomotor apraxia.

## Introduction

DNA single-strand breaks (SSBs) are amongst the most frequent lesions arising in cells, the threat of which is illustrated by the existence of six human genetic diseases in which one of four DNA single-strand repair proteins are mutated (Caldecott, 2022). These diseases are associated with neurodevelopmental and/or progressive neurodegeneration, and most commonly with cerebellar ataxia, oculomotor apraxia, and sensory neuropathy. SSBs arise from multiple sources including directly from attack of the sugar-phosphate backbone by reactive oxygen species, as obligate intermediates of DNA base excision repair, and as a consequence of the abortive activity of topoisomerases such as topoisomerase 1 (TOP1) (Caldecott, 2022).

The repair of SSBs is a multistep process in which SSBs are detected by PARP1 and/or PARP2, resulting in the activation and auto-modification of these sensor proteins with poly (ADP-ribose) at the site of the break, and likely also trans-modification of other proteins such as histones (Pandey & Black, 2021; Longarini & Matic, 2022; Huang & Kraus, 2022; Azarm & Smith, 2020). Poly (ADP-ribose) is a highly negatively-charged linear and/or branched polymer of up to several hundred ADP-ribose units that can relax chromatin compaction and accelerate DNA repair. Poly (ADP-ribose) also recruits XRCC1 protein complexes that contain the enzymes necessary for SSB repair such as TDP1, Aprataxin, PNKP, and DNA ligase III (LIG3). Notably, with the current exception of LIG3, mutations in all of these proteins are associated with hereditary neurodegenerative disease (Caldecott, 2022). In contrast, hereditary mutations in either of the SSB sensor proteins PARP1 or PARP2 in human disease have not been reported. Here, we present the first description of human hereditary disease in which PARP1 is mutated. We show that this disease is associated with reduced cellular levels of both ADP-ribosylation and DNA single-strand break repair (SSBR), and consistent with this that it is accompanied by progressive cerebellar ataxia and oculomotor apraxia.

## Results

Clinical analysis of a young girl (5-10 yr old) with childhood-onset neurodegeneration revealed rapidly progressive cerebellar ataxia with dystonia/parkinsonism and oculomotor dyspraxia (a detailed case report is available from the authors on appropriate request). Whilst an initial brain MRI when the proband was less than 5 yr old was normal, subsequent examinations 2 yr and 4 yr later revealed progressive cerebellar atrophy (Fig.1A). Exome sequencing identified a novel homozygous stop-gain variant in *PARP1* (c.384T>A; p.Cys128Ter) within a ∼21.1 Mb region of homozygosity on chromosome 1q41-43. Both parents were heterozygous for this variant (Fig.1B). This biallelic mutation is predicted to truncate PARP1 at the beginning of the second zinc finger, resulting in expression of a truncated protein of 127 amino acids encoding only the first zinc finger (Fig.1B) or, alternatively, in the absence of a truncated protein due to nonsense-mediated mRNA decay and/or protein instability. Consistent with the latter, we failed to detect PARP1 polypeptide in primary patient-derived fibroblasts using two different anti-PARP1 antibodies, one of which was raised against the amino terminal 300 amino acids (Fig.1C). This was in contrast to PARP2, which was present in patient-derived fibroblasts at levels similar to the parental cells (Fig.1C).

**Figure 1.**
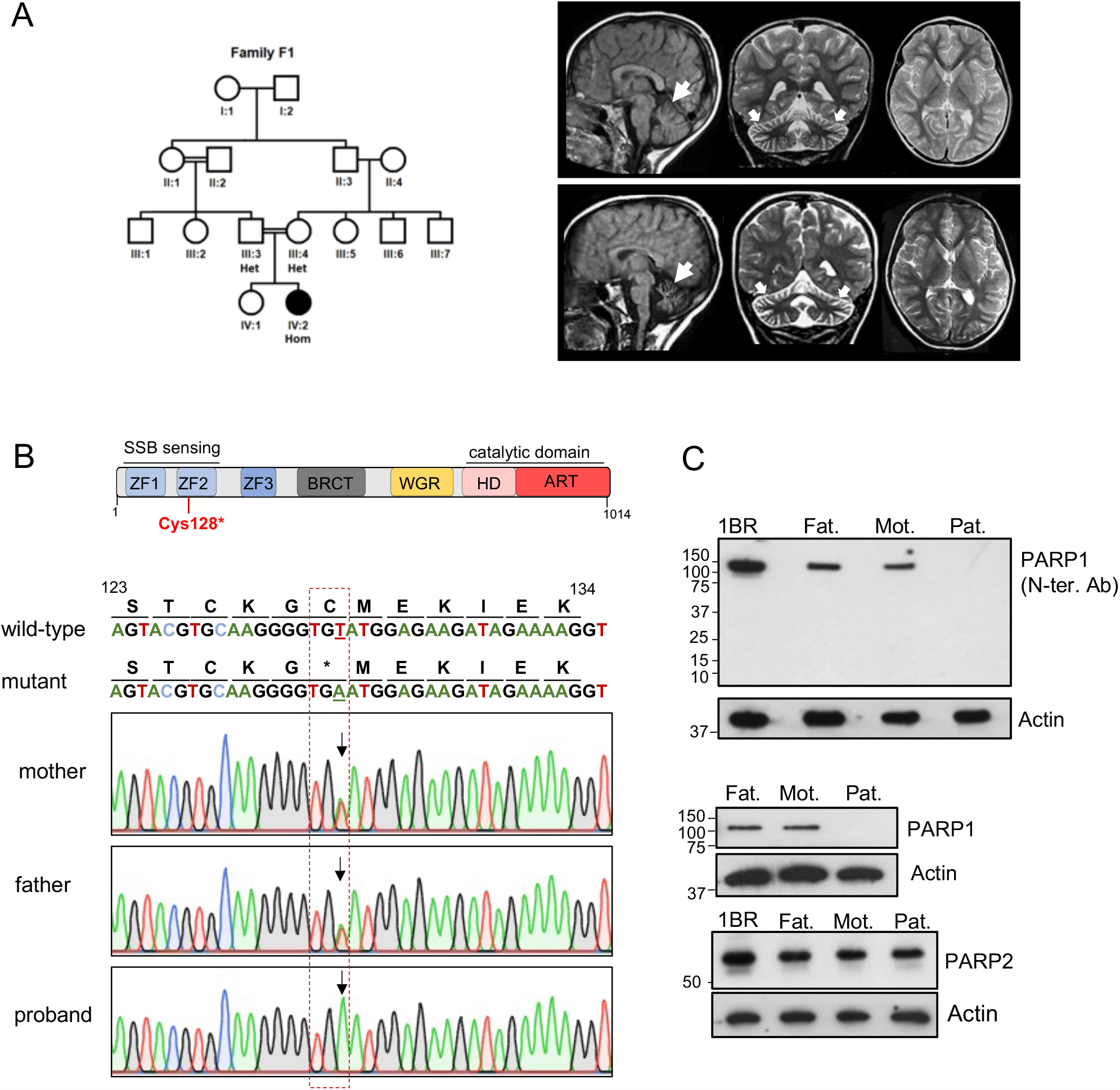
A patient with neurodegenerative disease and biallelic mutations in PARP1. **(A)**, Family tree (*left*) and MRI images (*right*) of the proband taken at < 5yr old and at 2 yr later, showing progressive atrophy of the cerebellum (arrows). **(B)**, *Top*, cartoon depicting the domain structure of PARP1, showing the position of the predicted stop codon introduced by the mutation c.384T>A (p.Cys128*). *Bottom*, confirmation of the biallelic *PARP1* mutation c.384T>A (p.Cys128*) in the affected proband, and heterozygosity in the unaffected parents, by Sanger sequencing. The sequences of wild type and mutated PARP1 alleles and protein are indicated. **(C)**, Western blots showing PARP1, PARP2, and actin protein levels in patient-derived primary fibroblasts from the unaffected proband (“Pat.”, patient), parents (“Mot.”, mother; “Fat.”, father), and unrelated normal human fibroblasts (1BR). The anti-PARP1 antibodies employed were raised against the amino-terminal 300 amino acids of recombinant human PARP1 (top blot) or a centrally located synthetic peptide (bottom blot).

To examine whether the patient-derived cells exhibited reduced levels of PARP activity, we measured levels of endogenous and genotoxin-induced poly (ADP-ribosylation). Levels of ADP-ribosylation induced by treatment with H_2_O_2_, a physiological source of oxidative SSBs, was reduced by ∼85% (Fig.2A). This low level of residual PARP activity is consistent with the previously reported contribution of PARP2 to DNA damage-induced ADP-ribosylation (Hanzlikova *et al*, 2017; Amé *et al*, 1999). Levels of endogenous ADP-ribosylation, detected primarily in S phase cells by incubation with an inhibitor of poly (ADP-ribose) glycohydrolase (PARG) to protect the nascent polymer (Hanzlikova *et al*, 2018), were also reduced (∼80%) in the patient cells (Fig.2B). Together, these data indicate that the biallelic patient mutations result in greatly reduced levels of PARP1 protein and activity.

**Figure 2.**
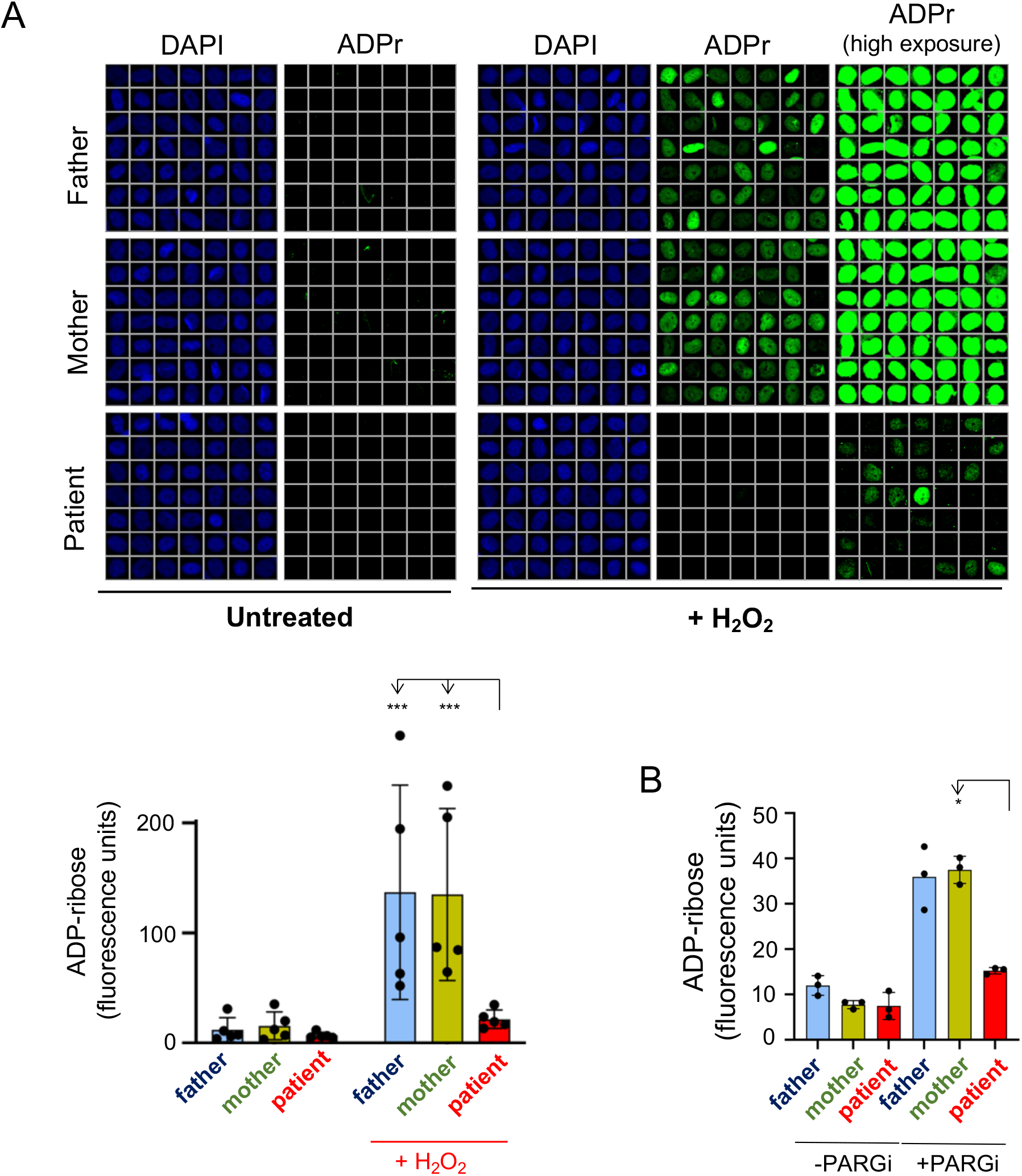
Greatly reduced levels of ADP-ribosylation in PARP1 patient-derived fibroblasts. **(A)**, representative single-cell galleries (*top*) and quantification (*bottom*) of nuclear ADP-ribose levels before and after treatment of cells with H_2_O_2_, from scanR high-content imaging. Quantified data are the mean (+/-1SD) ADP-ribose fluorescence from five independent experiments, with >1000 cells scored per sample per experiment. **(B)**, Quantification of endogenous ADP-ribose levels in S phase cells following, where indicated, incubation with PARG inhibitor (+PARGi) for 60 min to preserve nascent poly (ADP-ribose). Data were quantified as described above, from three independent experiments. Only significant differences are shown (*p≤0.05; ***p≤0.001), determined by 2-way ANOVA with Sidak’s multiple comparisons test.

Next, we examined whether the biallelic PARP1 mutations resulted in reduced rates of SSBR; one of the primary DNA repair pathways promoted by PARP1 (Caldecott, 2022). We treated cells with H_2_O_2_ and measured the induction and subsequent decline of SSBs using alkaline comet assays. Whereas oxidative SSBs were induced at similar levels in both parental and patient cells, the rate at which they declined was significantly reduced in the latter, with SSBR complete in parental cells after 15 minutes but still not complete in patient cells at the end of the experiment (Fig.3A). A similar defect was observed following treatment of the patient-derived fibroblasts with the alkylating agent MMS, which induces SSBs as obligate intermediates of DNA base excision repair (BER) (Fig.3B). For these experiments, we incubated the cells with MMS for 30 minutes and then measured the steady-state level of BER-associated DNA breaks. The median steady-state level of SSBs was increased ∼2.5-fold in the patient cells under these conditions when compared to the parental cells (Fig.3B), a phenotype typical of cells with inefficient SSBR (Taylor *et al*, 2002; Demin *et al*, 2021). Finally, we observed a similar defect following treatment of cells with camptothecin (CPT) for 30 minutes (Fig.3C), which induces SSBs by triggering the abortive activity of topoisomerase 1 (TOP1)(El-Khamisy *et al*, 2005). Such SSBs are implicated in a range of diseases associated with loss of efficient SSBR and cerebellar ataxia (El-Khamisy *et al*, 2005; Katyal *et al*, 2014; Hoch *et al*, 2017). Under the conditions employed, the median steady-state level of TOP1-SSBs was increased ∼4-fold in the PARP1 patient cells, when compared to parental cells (Fig.3C).

**Figure 3.**
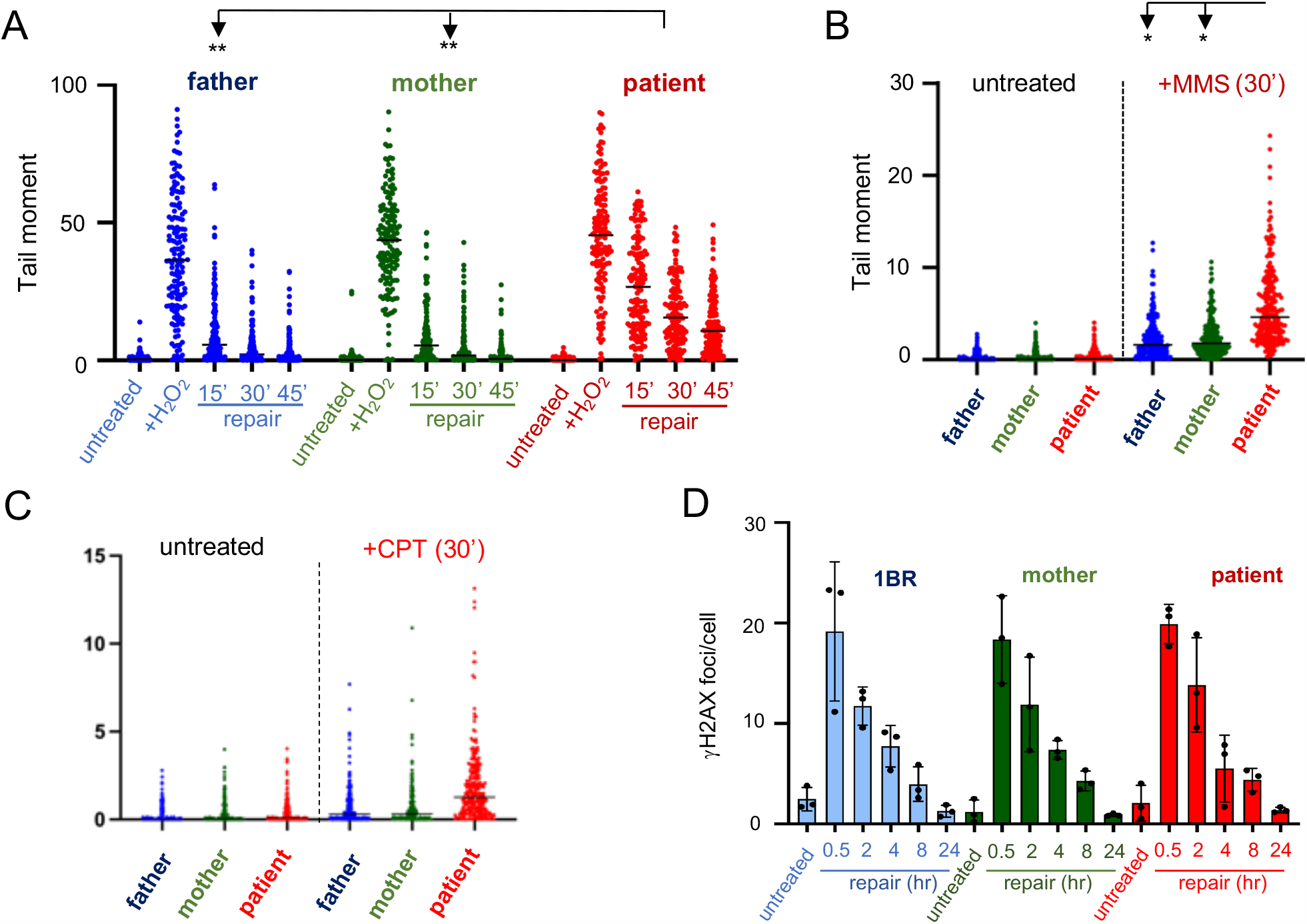
Reduced rates of DNA single-strand break repair in PARP1 patient-derived fibroblasts. **(A)**, DNA strand breaks quantified by alkaline comet assays in the indicated primary human fibroblasts before (untreated) and immediately after treatment or not with 25 μM H_2_O_2_ for 10 min on ice, and at the indicated times following drug treatment. Data plotted are the individual comet tail moments (an arbitrary measure of DNA strand breakage) combined from three independent experiments (50 cells per sample per experiment). **(B)**, DNA strand breaks quantified as above following treatment or not with 0.05 mg/ml MMS for 30 min. **(C)**, DNA strand breaks quantified as above following treatment or not with 10μM camptothecin (CPT) for 30 min as described above. For the above analyses, only significant differences are shown (*p≤0.05; **p≤0.01), determined by 2-way ANOVA with Tukey’s multiple comparisons test. **(D)**, DNA double-strand breaks quantified by measuring the number of γH2AX foci per cell before and immediately after γ-irradiation (2Gy), and at the indicated times (hrs) following irradiation. No significant differences between cell lines were detected, using one-way ANOVA with Holm-Sidak’s multiple comparisons test.

Next, we measured the rate of DNA double-strand break repair (DSB) in the parental and patient-derived cells, because PARP1 is implicated in the repair of DSBs by non-homologous end-joining (NHEJ)(Huang & Kraus, 2022). To measure DSBs, we exposed cells to γ-irradiation and quantified the number of nuclear γH2AX immunofoci, which are an indirect but robust measure of DSBs following ionising radiation (Löbrich *et al*, 2010). Exposure to of γ-rays (2 Gy) induced similar levels of γH2AX foci in parental and patient cells, and in unrelated 1BR normal human fibroblasts. More importantly, the rate at which γH2AX foci declined following irradiation was similar in all of the cell lines, suggesting that the rate of DSB repair was normal in the PARP1 patient-derived cells.

Finally, we examined whether the reduced rates of SSBR in patient-derived cells resulted in hypersensitivity to genotoxins. Indeed, the patient-derived fibroblasts were hypersensitive to CPT (Fig.4A), consistent with their reduced rate of repair of TOP1-induced SSBs. In contrast, we did not exhibit significant hypersensitivity to ionising radiation (Fig.4B), the cytotoxicity of which is driven primarily by DSBs.

**Figure 4.**
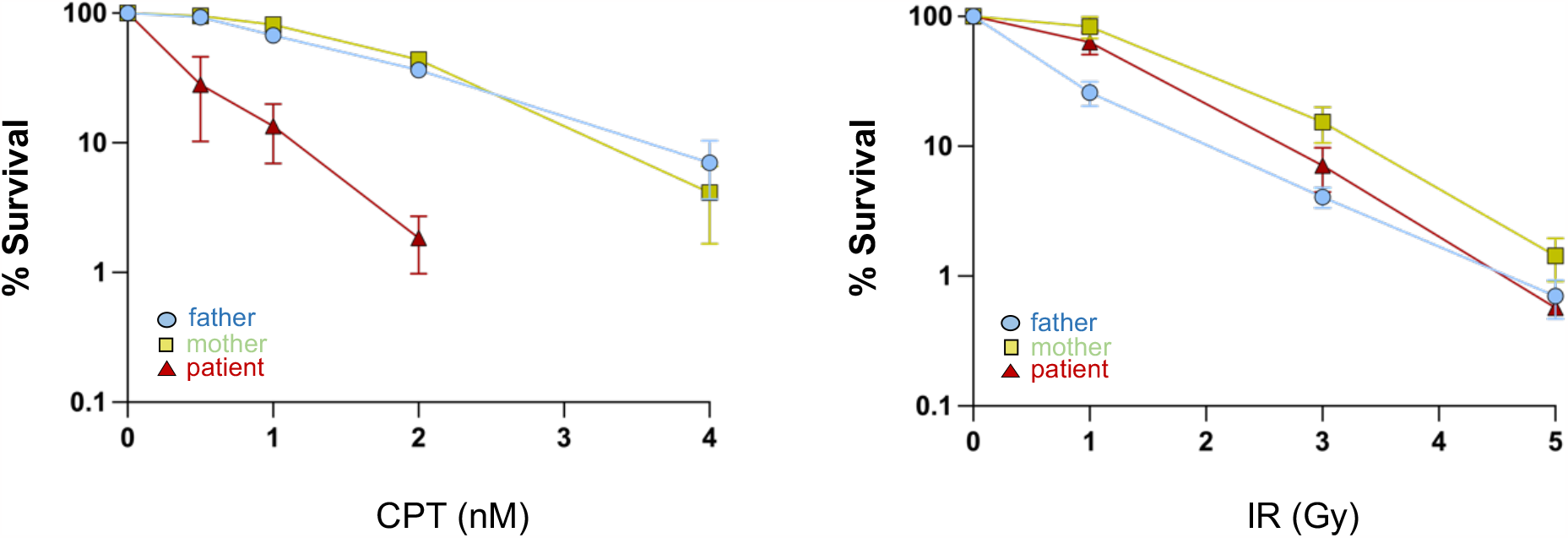
Hypersensitivity to TOP1-associated SSBs induced by camptothecin in PARP1 patient-derived fibroblasts. Primary human fibroblasts plated in 10cm dishes were treated with the indicated doses of camptothecin (CPT, *left*) or γ-rays (right) and after 12-14 days cells were fixed, stained, and surviving colonies (>50 cells) were counted. Data are the mean (+/SD) of three independent experiments.

## Discussion

Defects in SSBR are associated with a spectrum of neurological pathologies, but most commonly with neurodegeneration typified by progressive cerebellar ataxia and oculomotor apraxia (Caldecott, 2022; Yoon & Caldecott, 2018; McKinnon & Caldecott, 2007). To date, all known SSBR-associated diseases result from mutations in proteins involved in DNA end processing, the stage of SSBR in which DNA termini are restored to the chemistry appropriate for DNA ligation. These diseases include the three neurodevelopmental and/or neurodegenerative diseases *microcephaly with early onset seizures* (MCSZ; OMIM 613402), *ataxia oculomotor apraxia-4* (AOA4; OMIM 616267) and *Charcot-Marie Tooth disease Type 2B2* (CMT2B2; OMIM 605589) arising from mutations in PNKP (Shen *et al*, 2010; Leal *et al*, 2018; Kalasova *et al*, 2020; Bras *et al*, 2015), and the two neurodegenerative diseases *spinocerebellar ataxia with axonal neuropathy-1* (SCAN1; OMIM 607250) and *ataxia oculomotor apraxia-1* (AOA1; OMIM 208920) arising from mutations in the proteins TDP1 or APTX, respectively (Takashima *et al*, 2002; Moreira *et al*, 2001; Date *et al*, 2001). These three proteins assemble into a multiprotein complex by interaction with the scaffold protein XRCC1, which itself is mutated in a fifth neurodegenerative disease, known as ataxia oculomotor apraxia-5 (spinocerebellar ataxia recessive 26 SCAR26/AOA5; OMIM 617633)(Hoch *et al*, 2017; O’Connor *et al*, 2018).

Importantly, PARP1 has not previously been implicated in hereditary neurological disease, despite the importance of this sensor protein for rapid recruitment of the XRCC1 protein complexes described above. A possible explanation for this has been the extensive enzymatic redundancy of PARP1 with PARP2 (Murcia *et al*, 2003; Hanzlikova *et al*, 2017). However, our data now suggest that, similar to other SSBR proteins, mutations in PARP1 are associated with progressive cerebellar ataxia and oculomotor apraxia. Whilst we have so far identified and characterised only one patient with biallelic PARP1 mutations, it is very likely that these mutations are the cause of the associated neuropathology. This is because we failed to identify any other putative pathogenic mutations in this patient by exome sequencing, and because the cerebellar ataxia and oculomotor apraxia observed in the current proband is very similar to that reported for other SSBR-defective diseases (Hoch *et al*, 2017; Kalasova *et al*, 2019, 2020; El-Khamisy *et al*, 2005). Indeed, the biallelic PARP mutations identified here reduced SSBR in the patient cells to rates similar to those observed in other SSBR-defective diseases.

The PARP1 biallelic mutations truncate PARP1 at the beginning of the second DNA strand-break sensing zinc finger domain, preventing both SSB binding and subsequent catalytic activity (Murcia *et al*, 1989; Eustermann *et al*, 2015; Sefer *et al*, 2022). Consistent with this, we detected only 10-20% residual poly (ADP-ribose) activity in patient-derived primary fibroblasts, a level similar to that observed previously in cell lines in which PARP1 has been deleted (Hanzlikova *et al*, 2017, 2018). We did not detect in the patient cells the truncated PARP1 peptide of 127 amino acids predicted by the biallelic mutations, suggesting that the peptide is unstable or is expressed at very low levels as a result of nonsense-mediated mRNA decay. Alternatively, the truncated peptide may lack the epitopes detected by the monoclonal antibodies employed here, though we note that one of the antibodies was raised against a 300 amino acid amino-terminal fragment of PARP1.

Whilst our data implicate PARP1 loss as a cause of neurodegenerative disease, PARP1 deletion rescues much of the neuropathology observed in a mouse model of XRCC1-defective disease (Hoch *et al*, 2017; Komulainen *et al*, 2021). The PARP1-dependent neuropathology in XRCC1-defective brain appears to reflect a key role for XRCC1 in preventing excessive SSB engagement and interference by PARP1 during DNA base excision repair (BER), which otherwise results in PARP1 ‘trapping’ on BER intermediates and much slower rates of BER (Adamowicz *et al*, 2021; Demin *et al*, 2021). In contrast, the disease reported here is most likely triggered by PARP1 loss of function, as a result of the slower repair of SSBs arising from oxidative stress and/or abortive topoisomerase activity (Caldecott *et al*, 2022).

## Materials and Methods

### Exome sequencing

To identify the pathogenic variant responsible for the disease in the patient, repeat expansion for FXN followed by exome sequencing was carried out at Centogene diagnostic lab as previously described and results came back negative. Further re-analysis of the sequencing data at Queen Square Genomic research laboratory, identified a novel homozygous stop-gain variant in *PARP1*(NM_001618.4), c.384T>A (p.Cys128Ter) encompassing ∼21.1 Mb region of homozygosity on chromosome 1q41-43. Both parents were carriers of the variants.

### Cell Culture

Primary fibroblasts derived from the proband and parents, and also unrelated 1BR fibroblasts, were maintained in Dulbecco’s minimal essential medium (DMEM) supplemented with 10-15% foetal calf serum (FCS), penicillin/streptomycin and L-glutamine/GlutaMAX. All cells were grown at 37°C in a low oxygen (3 %) incubator. For the purpose of reagent requests, the fibroblast cell lines employed here are denoted ION-1041 (Father), ION-1042 (Mother), and ION-1043 (Affected child), and the unrelated control is 1BR. Please note that these names are anonymised catalogue names and unrelated to individual identities.

### Clonogenic survival assays

Cellular sensitivity was assessed by colony survival analysis. Patient fibroblasts were either irradiated using a ^137^Cs γ-ray source at a dose rate of 0.09 Gy s^-1^ or treated continuously with Camptothecin (CPT) at the doses stated. After treatment, fibroblasts were plated onto lethally-irradiated feeder cells prepared 24 h earlier, and colony staining and counting via Methylene Blue carried out three weeks later.

### Western blotting

Cells were lysed directly in SDS-PAGE sample buffer, heated for 10 min at 95°C and subjected to SDS-PAGE followed by protein transfer onto nitrocellulose membrane and western blotting with the indicated antibodies. The antibodies employed were the anti-PARP1 mouse monoclonal B10 (Santa Cruz sc-74470, used at 1/1000 dilution) raised against a recombinant protein encoding the N-terminal 300 amino acids of human PARP1, and the anti-PARP1 rabbit monoclonal 46D11 (Cell Signalling Technologies #9532, used at 1/1000 dilution) raised against a synthetic peptide spanning the residue Gly623 near the middle of human PARP1, anti-PARP2 rabbit polyclonal (Active Motif #39743, used at 1/1000 dilution), anti-β-Actin mouse monoclonal (Proteintech #66009-1-Ig, used at 1/5000), HRP-conjugated rabbit anti-mouse secondary antibody (Dako P0260, used at 1/10000 dilution), and goat HRP-conjugated anti-rabbit secondary antibody (Dako P0448, used at 1/10000 dlution).

### Alkaline comet assays

Where indicated, cells were treated in PBS with 25μM hydrogen peroxide (H_2_O_2_) on ice for 10 min, or with 0.05 mg/ml MMS or 10μM CPT for 30 min at 37°C in growth medium. Following treatment, cells were washed once in ice-cold PBS and resuspended in 0.4 mL PBS. Aliquots (0.15 ml) of the cell suspension were mixed with an equal volume of low-gelling temperature agarose (at 42°C), spread on agarose pre-coated frosted slides, and the gel suspension allowed to set on ice. The cells were then lysed in alkaline lysis buffer (2.5M NaCl, 100 mM EDTA, 10 mM Tris-Cl, 1% v/v DMSO, 1% v/v Triton X-100, pH 10) at 4°C for 60 min. Samples were pre-incubated for 45 min in electrophoresis buffer (50mM NaOH, 1mM EDTA, 1% DMSO, pH13) and then fractionated at 12V for 25 min, at 4°C. Slides were neutralized in 0.4M Tris-Cl pH 7.4 prior to staining in PBS containing SYBR Green (Sigma, 1:10000 dilution) and 0.04 mg/ml p-Phenyledenediamine dihydrochloride anti-fade (Fisher 417481000). Comet tail moments (an arbitrary measure of DNA strand breaks) from 50-100 cells per sample were blinded and scored using automated Comet Assay IV software (Perceptive Instruments, UK).

### DSB repair assays

Cells were grown on coverslips until confluent and irradiated with 2 Gy γ-radiation. After treatment, cells were rinsed and fixed for 10 min in PBS containing 4% paraformaldehyde at the indicated time points. Cells were permeabilized (20 min in PBS containing 0.2% Triton X-100), blocked (1 hour in PBS-5% BSA), and incubated with anti-γH2AX (Millipore #05-636, used at 1:2500 dilution) for 3 hours in PBS containing 5% BSA. Cells were then washed (3×5 minutes in PBS containing 0.1%Tween-20), incubated for 1 hour with goat anti-mouse Alexa Fluor 488 conjugated secondary antibody (1:1000, 5% BSA), and washed again as described above. Cells were counterstained with DAPI and mounted in VECTASHIELD.

### ADP-Ribosylation Assays

Cells grown on coverslips were treated with either 100 μM H_2_O_2_ in PBS on ice for 10 min or 10 μM PARG Inhibitor (PDD0017273) in complete culture media for 1 hour at 37°C degrees. After treatment, cells were rinsed twice with PBS and incubated with pre-extraction buffer (25 mM HEPES pH 7.4, 50 mM NaCl, 1 mM EDTA, 3 mM MgCl2, 0.3 M sucrose, 0.5% Triton X-100) for 5 min on ice, and then fixed with cold 4% formaldehyde for 15 min and permeabilized using PBS containing 0.5% Triton X-100. After blocking (1 hour in PBS containing 5% BSA), cells were incubated with the anti-poly/mono ADP-ribose nanobody Macro(Af1521)^MG^-Fc at 1/10000 (for cells treated with H_2_O_2_), or with anti-poly/mono ADP-ribose rabbit monoclonal antibody E6F6A (Cell Signalling Technology #83732) at 1/500 dilution for 2 hours in PBS containing 5% BSA (for cells incubated with PARG inhibitor). Cells were then washed (3×5 min in PBS containing 0.1%Tween-20), incubated for 1 hour with the corresponding Alexa Fluor 488 goat anti-rabbit or anti-mouse secondary antibody (diluted at 1:1000 in 5% BSA), and washed again. Finally, cells were counterstained with DAPI and mounted in VECTASHIELD. Numbers of γH2AX foci per cell and poly/mono ADP-ribose immunofluorescence were quantified using an Olympus ScanR high-content imaging system with ScanR Image Acquisition and Analysis Software.

## Data Availability

All data present in the manuscript are available on reasonable request, following publication.

## Acknowledgements

We thank the parents and proband for their consent to participate in and publish this study.

